# Estimating the state of the Covid-19 epidemic in France using a non-Markovian model

**DOI:** 10.1101/2020.06.27.20141671

**Authors:** Raphaël Forien, Guodong Pang, Étienne Pardoux

## Abstract

In this paper, we use a deterministic non-Markovian epidemic model to estimate the state of the Covid-19 epidemic in France. This model allows us to consider realistic distributions for the exposed and infectious periods in a SEIR model, contrary to standard ODE models which only consider exponentially distributed exposed and infectious periods. We present theoretical results linking the (unobserved) parameters of the model to various quantities which are more easily measured during the early stages of an epidemic. We also stress the main quantitative differences between the non-Markovian and the Markovian (ODE) model. We then apply these results to estimate the state of the Covid-19 epidemic in France by analyzing three regions: the Paris region, the northeast regions and the rest of the country, based on current knowledge on the infection fatality ratio and the exposed and infectious periods distributions for Covid-19. Our analysis is based on the hospital data published daily by Santé Publique France (daily hospital admissions, intensive care unit admissions and hospital deaths).

## Introduction

At the beginning of an epidemic outbreak, some quantities are easier to observe and report than others. For example the number of hospital deaths related to Covid-19 has been precisely and regularly reported in several countries. Another well documented quantity is the doubling time of the number of cases (which coincides with that of the number of deaths, as we shall explain). On the other hand, some quantities of interest are very hard to directly measure or to estimate: the actual number of infected individuals, the true death rate of the disease, and most notably the now famous reproduction number *R*_0_, corresponding to the average number of individuals infected by a single infectious individual in a fully susceptible population.

This paper presents a theoretical study of how the “observable” quantities relate to the “unobserved” ones, under a very general epidemic model recently developed in [PP20], and an application of these results to attempt to predict the state of the Covid-19 epidemic in France throughout its different stages.

The first basic idea is to estimate an exponential growth rate from the available data (the number of hospital admissions or the number of deaths occurring in hospitals), and then to relate it to the parameters of a model of the evolution of the Covid–19 epidemic. Our approach is modeled upon the approach of Tom Britton in two recent preprints [Bri20b], [Bri20a]. Note that the exponential growth rate can be negative, which was the case in particular in France during the lockdown period.

The exponential growth rate is the most natural parameter which can be extracted from the data. It equals log(2) divided by the doubling time, i.e. the number of days necessary for the number of cases (or deaths) to double, a notion that everyone listening to the news at the time of the rise of the epidemic has heard of, and can easily understand. The other important parameters of the model, including the famous basic reproduction number *R*_0_ (the mean number of individuals whom an infectious individual infects before recovering, at the start of the epidemic – that is while essentially everyone that around him/her is susceptible), can be computed from the exponential growth rate, and some other parameters of the model.

Our approach has two specific features. First, we shall carefully make explicit which parameters are needed to compute the quantities of interest, and we shall describe as precisely as possible how the the uncertainty about their value influences our results. The second aspect is that we shall use nonconventional models. The classical SIR and SEIR ODE models are law of large numbers limits of stochastic Markov SIR or SEIR models [BP19]. For those models to be Markovian, it is necessary that the infectious periods in the case of the SIR model of the various individuals in the population be i.i.d. copies of an exponential random variable (resp. the pair exposed period, infectious period in the SEIR model be i.i.d. copies of a pair of independent exponential random variables). However, in the case of Covid–19, like in most infectious diseases, the assumption that those durations follow an exponential distribution is completely unrealistic.

Recently, the last two authors of this paper have described the law of large numbers limit model of non–Markovian stochastic epidemic models with arbitrary distribution for the infectious period (resp. for the pair exposed and infectious period), see [PP20]. This type of deterministic models is an integral equation of Volterra type, of the same dimension as the classical ODE model. In particular, it is not much more complicated to simulate and compute. The differences between our model and the usual “Markov” model are not so much in the large time behavior, but rather in the transient short term evolution, as was recently observed in [SRE^+^20], who study a related model. We note that since the start of the epidemic, knowledge about the Covid–19 disease has substantially increased. Our original approach allows us to choose distributions which reflect closely the current knowledge about the disease.

Note that at least one other work has used a similar extended model to analyze the Covid–19 epidemic. In [FKK20], the authors use ODEs with delays, which correspond to our model with deterministic exposed and infectious periods. On the other hand [GAA^+^20], following the approach initiated by Kermack and McKendrick to analyze the plague epidemic in Mumbai in 1905–6 [KM27], uses a transport PDE SEIR model, where the rate of infection by an infectious individual depends upon the time since infection, and the rate at which exposed (resp. infectious) individuals become infectious (resp. removed) also depends on the time since infection. The results of the present paper are comparable to those in [GAA^+^20], where the analysis and the treated data concern the various *départements* of Î le de France, while we compare Î le de France, the North–East of France, and the rest of the country.

Models similar to our non-Markovian SEIR epidemic model appear in several papers (see, e.g., [AW80, YF10]) and in section 4.5 of the recent book [BCCF19]. However, our model seems to be more general than those who appeared earlier, in particular regarding the initial condition and allowing correlated exposed and infectious periods, and as far as we can tell its rigorous interpretation as the functional law of large numbers limit of an individual based stochastic model established in [PP20] is new. This interpretation was crucial to suggest the reduction of the dimension of the model which we shall describe below. As a matter of fact, our formula for *R*_0_ may be considered as a particular case of a formula which appears on page 141 of [BCCF19].

The paper is organized as follows. In a first section called “Methods”, we describe our model and the methodology to extract the parameters of our model from the available data. In a second section called “Results”, we describe the results we obtained by applying our method to the Covid-19 epidemic in France. The last section is a discussion of the conclusions of our work. Finally, an appendix contains the mathematical proof of one crucial result, which relates the exponential growth rate to the various quantities in our model.

## 1 Methods

### 1.1 Our Covid–19 non–Markovian SEIR epidemic model

Assume that each newly infected individual in the population becomes infectious after a random time *ε* during which he or she is “exposed” and stops being infectious after a random time ℐ, after which he or she does not infect anyone any more, and also cannot be infected (either as a result of acquired immunity, isolation or death). We assume that infectious individuals attempt to infect other individuals (chosen uniformly from the population) at rate *λ*(*t*), where *t* is the current time (the dependence on *t* of the contact rate *λ* is used to reflect the effect of containment measures such as lockdown). As a result, the process of newly infected individuals up to time *t* can be represented as

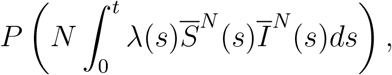

where *P* (*t*) is a standard Poisson process, *N* is the size of the population which is assumed to be fixed, 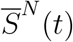 (resp. *Ī*^*N*^ (*t*)) is the proportion of susceptible (resp. infectious) individuals in the population at time *t*.

Assume that the initial proportions of susceptible, exposed, infectious and removed individuals in the population are given by 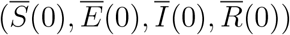. We also assume that the individuals who are initially exposed or infectious have a different distribution of sojourn times in the intermediary states (reflecting the fact that they have been infected at some time in the past, before the initial time *t* = 0). Thus let (*ε*_0_, ℐ_0_) denote a random variable distributed as the time an individual who is initially exposed stays exposed (*ε*_0_) and then infectious (ℐ_0_), and let ℐ_1_ be distributed as the time an individual who is initially infectious stays infectious. Note that usually ℐ_0_ and *I* have the same distribution for those who are initially exposed, while the remaining infectious period *I*_1_ may have a different distribution. Similarly, the remaining exposed period *ε*_0_ may have a different distribution from that of *ε*.

Define

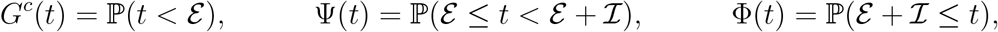

as well as

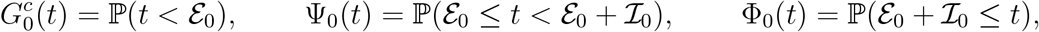

and

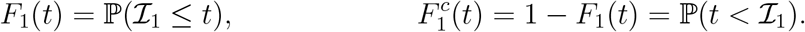

Let 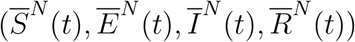 denote the proportion of susceptible, exposed, infectious and removed individuals at time *t* in the population, where *N* is the total size of the population, which is assumed to be fixed throughout the epidemic (the deaths due to the epidemic are counted among the “removed”). We assume that the durations (*ε*_*i*_, ℐ_*i*_) of all the individuals infected during the epidemic are i.i.d. (for each *i, ε*_*i*_ and ℐ_*i*_ may be correlated).

**Definition 1**. *The deterministic non-Markovian SEIR model is the solution of the set of integral equations:*

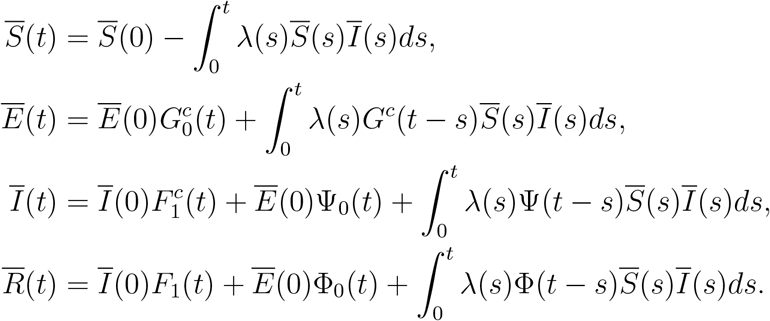

Theorem 3.1 from [PP20] states that, under a very weak assumption on the joint distribution of (*ε*, ℐ), the unique solution of the above non-Markovian SEIR integral equations is the law of large numbers limit of the above described model. More precisely, as *N* → ∞, 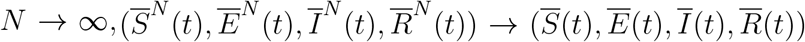 in probability, locally uniformly in *t*. We call this model non–Markovian since the stochastic process 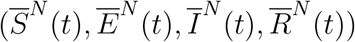 is not a Markov process in general, because its future evolution depends not only upon its present value, but also upon how long the exposed and infectious individuals have already been in the corresponding compartment. For the model to be Markovian, it is necessary that the random variables ℐand ℐ be independent and have exponential distributions. In that case, the integral equation SEIR model reduces to the following ODE model

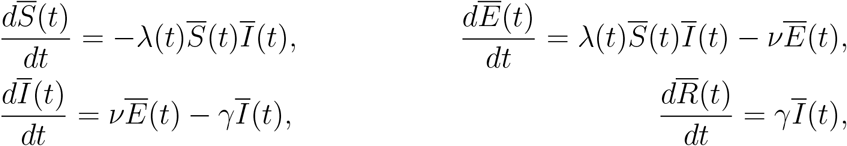

where *ν* (resp. *γ*) is the parameter of the exponential law of *ε* (resp. ℐ), i.e., 𝔼(*ε*) = *ν*^−1^, 𝔼(ℐ) *γ*^−1^ Note that in the Markovian case, the convergence 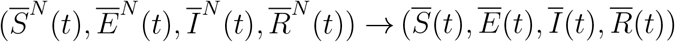 as *N* → ∞ is a classical result, see, e.g., [BP19] for a recent account.The above ODE model is the one which is used by almost all papers dealing with epidemic models, in particular the Covid–19 models, with the exception, to our knowledge, of [FKK20], where the case of fixed *ε* and ℐ is considered and [GAA^+^20, SRE^+^20]. It is fair to say that the exponential distributions are very poor models for the laws of *ε* and ℐ, and it may seem strange that the above ODE model is so widely used, while it is based on unrealistic assumptions. While the large time behavior of the model (e.g., the endemic equilibrium in the case of a SIS or a SIRS model, see Remarks 2.8 and 3.5 in [PP20]) depend only on the expectation of the random vector (*ε*, ℐ), and are the same in a Markov and a non-Markov model with identical expected infectious periods, as we shall see below, the transient behavior of the model depends much more on the details of the law of (*ε*, ℐ).

Let us now explain the specific model (with two variants) which we will use for the Covid–19 epidemic. In our model, the two random variables *ε* and ℐ will be independent. The random variable *ε* will be either fixed (3 days), or else will have a distribution with a density whose support is the interval [2, 4]. Concerning the random variable ℐ, we assume a bimodal law with support in [3, 5] ⋃ [8, 12] (details in Subsection 1.4).

The idea behind that type of law is as follows. Our model is close to the SEIRU model of [LMSW20], which thanks to the flexibility of our class of models we are able to simplify. Indeed, in that paper the individuals are first exposed (state E), then infectious (state I) but without symptoms, and at the end of the I period, a fraction of the individuals are isolated quickly after the onset of symptoms (state R as “reported”), either because they are admitted to hospital or because they self-isolate at home, while the other individuals are not isolated (state U as “unreported”), either because they are unable to do so or because they have light or no symptoms. While in the R class, the individuals are isolated and they do not infect susceptibles any more. For that reason, we identify the “reported” class with part of the “recovered” class, thus reconciling the two meanings of R. Finally, some of the individuals are infectious while they are in the I class only, while others are infectious in both I and U classes. In our model, being in the U class is considered as staying longer in the I class (see Figure 1). This is the motivation for the bimodal distribution of the law of ℐ.

**Figure 1:**
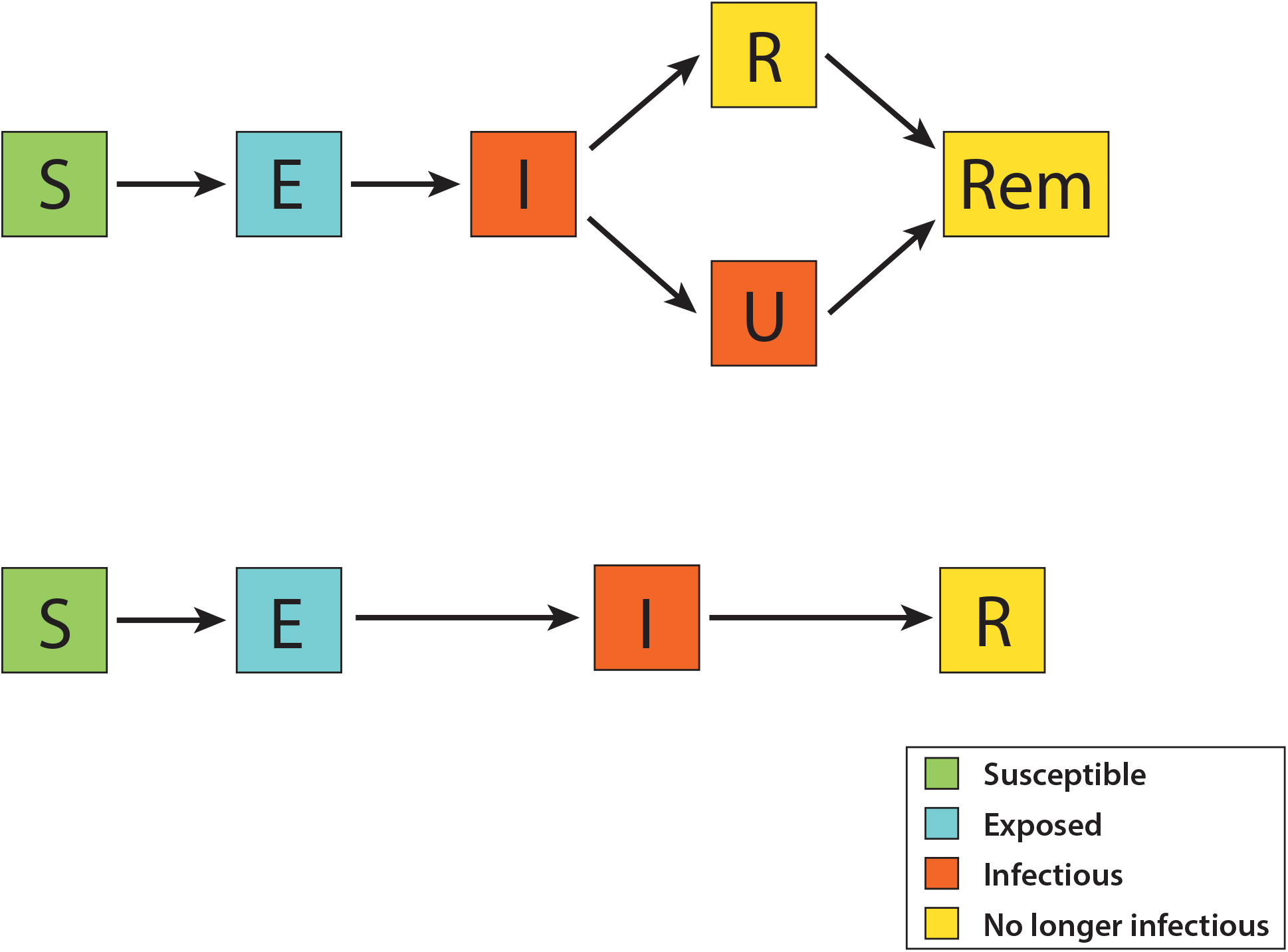
Flow chart of the SEIRU model of [LMSW20] and the equivalent SEIR non-Markovian model. In the latter, the two infectious compartments (I and U) are merged into one compartment, but the sojourn time ℐ of the infectious compartment has a bimodal distribution corresponding to the two subpopulations of reported and unreported individuals. Note that R (reported) and Rem (removed) have been merged into a unique compartment.

#### Remark 2.

*Our model lacks one important feature which is widely believed to affect the dynamics of the Covid-19 epidemic, namely age structure. Our main motivation for neglecting age structure is to keep the model mathematically tractable and to reduce the number of parameters. Needless to say, more realistic predictions could be obtained by using a structured version of this model, provided all the parameters can be correctly estimated*.

### 1.2 The non–Markovian model during the early phase of the epidemic

During the early phase of the epidemic, the cumulative number of infected individuals remains small compared to the total size of the population. As a result, 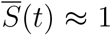 during this phase. Letting (*E*(*t*), *I*(*t*), *R*(*t*)) denote the absolute numbers of exposed, infectious and removed individuals during this phase, *i.e*.,

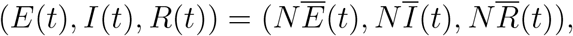

and assuming that *λ* is constant during this phase, the non-Markovian SEIR model reduces to

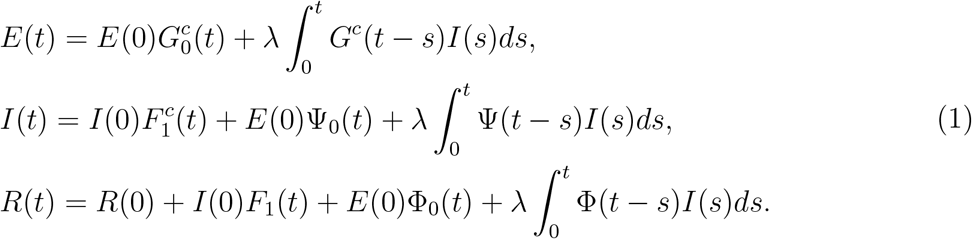

The initial state of an epidemic is seldom known, and the Covid-19 epidemic is no exception. However, it is always the case that, once sufficiently many individuals have been infected, the cumulative number of infected individuals grows exponentially with some rate *ρ* > 0. We thus look for solutions to

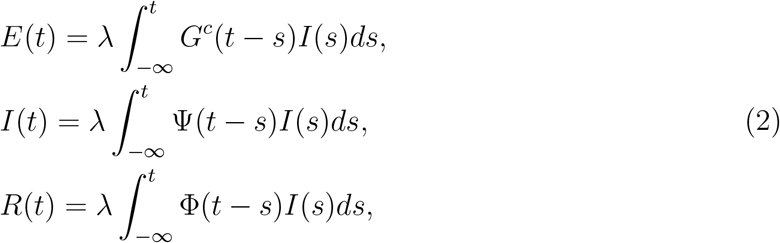

for *t* ∈ ℝ which are of the form

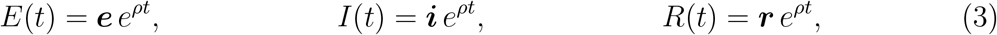

with ***e*** + ***i*** + ***r*** = 1. These equations started from −∞ can be seen as modeling an epidemic which has been growing for a very long time from an infinitesimal proportion of non-susceptible individuals.

#### Proposition 3.

(*i*) *If ρ* > 0, *the system* (2) *admits solutions of the form* (3) *for all t* ∈ ℝ

*if*

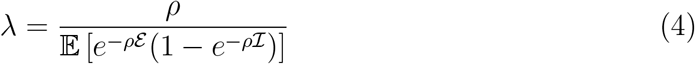

*and with*

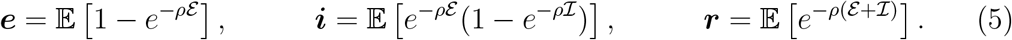

*Moreover, if* Θ *is an independent exponential variable with parameter ρ, then* (3) *also solves* (1) *for all t* ≥ 0 *with*

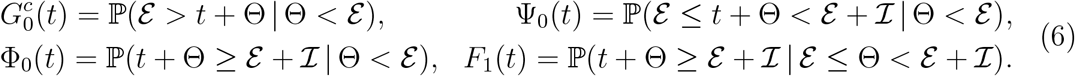

(*ii*) *If ρ* < 0, *then* (*E*(*t*), *I*(*t*)) = (e*e*^*ρt*^, i*e*^*ρt*^) *solves the first two lines of* (2) *for all* ∈ ℝ*if λ and ρ satisfy* (4) *and if*

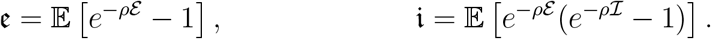

The fact that a solution of the form *R*(*t*) = ***r*** *e*^*ρt*^ only exists for positive *ρ* should not come as a surprise, since *t ↦ R*(*t*) is non-decreasing. Also note that we can rewrite (5) with the help of the variable Θ as follows,

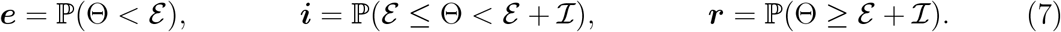

The proof of Proposition 3 is postponed to Appendix A.

#### Corollary 4.

*The basic reproduction number R*_0_, *defined as the mean number of secondary infections caused by a single infectious individual in a fully susceptible population, is linked to the initial growth rate of the cumulative number of infected individuals ρ by the relation*

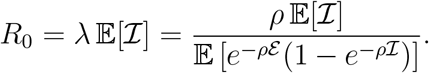

*This formula is valid in both cases ρ* > 0 *and ρ* < 0.

#### Remark 5.

*Note that if* (*ε*, ℐ) *are two independent exponential random variables with parameters ν* > 0 *and γ* > 0, *the equations of Definition 1 coincide with the law of large numbers limit of the Markovian SEIR epidemic model* [BP19]. *In this case, Proposition 3 agrees with what is already known about Markovian epidemic models. In particular, in the case ρ* > 0,

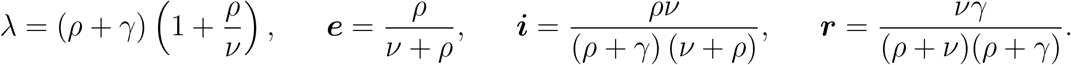

*The equivalent formulas for the SIR model* (*i.e*., *with ε* = 0) *can be obtained by letting ν* → ∞, *in which case we see that ρ* = *γ*(*R*_0_ − 1), *as in formula* (*1*) *in* [Bri20b].

*If, however, we assume that the variables and are constant ε and* ℐ *equal to* (*t*_*e*_, *t*_*i*_), *the equations of Definition 1 can be seen as delay equations. In this case, Proposition 3 still applies, and the expectations in* (4) *and* (5) *can be omitted, leading to the relation*

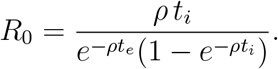

*As a result, we see that, for the same growth rate ρ, the contact rate and the relative proportions of exposed, infectious and removed individuals vary depending on the distribution of the sojourn times* (*ε*, ℐ). *In particular, the growth rate ρ cannot be used to accurately determine the value of R*_0_ = *λ* 𝔼[ℐ] *if one lacks information about the distribution of* (*ε*, ℐ) (*see Figure 2*).

**Figure 2:**
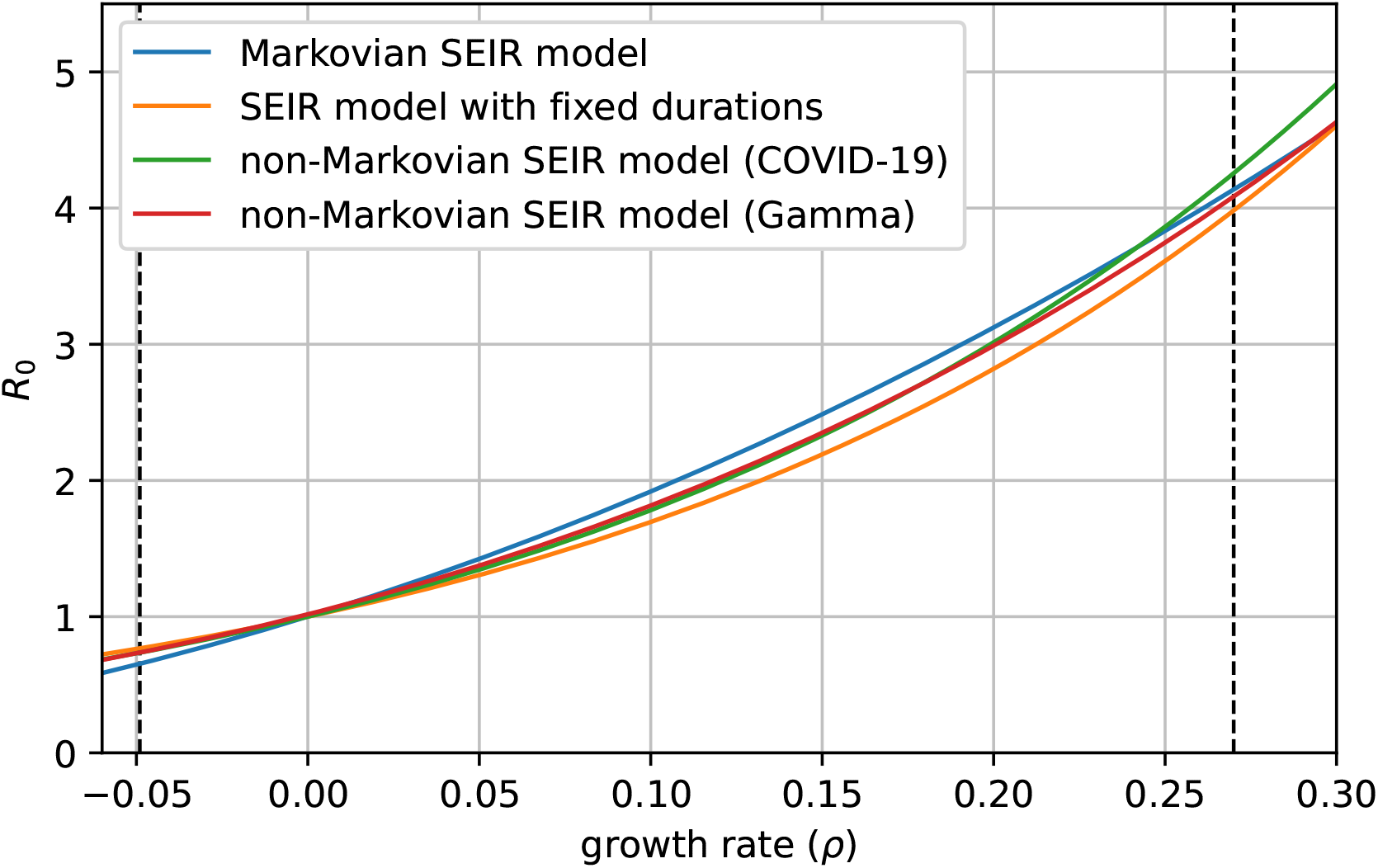
Value of *R*_0_ as a function of the growth rate *ρ* for different distributions of the exposed and infectious periods (*ε*, ℐ). Four types of distributions are displayed: exponential (corresponding to the Markovian SEIR model), fixed, bimodal distribution mimicking Covid-19 (see Subsection 1.4) and Gamma distribution. All distributions have a mean exposed time of 3 days and a mean infectious time of 4.8 days, corresponding to a proportion of reported individuals of 0.8. The two dashed vertical lines show the growth rates of the Covid-19 epidemic in mainland France before and during lockdown.

### 1.3 Changing the contact rate during the early phase of the epidemic

In France, as in many countries around the globe, strict lockdown measures were put into place at a fairly early stage of the COVID-19 epidemic. Assuming that, at this stage, the proportion of non-susceptible individuals remains negligible, the non-Markovian model reduces to

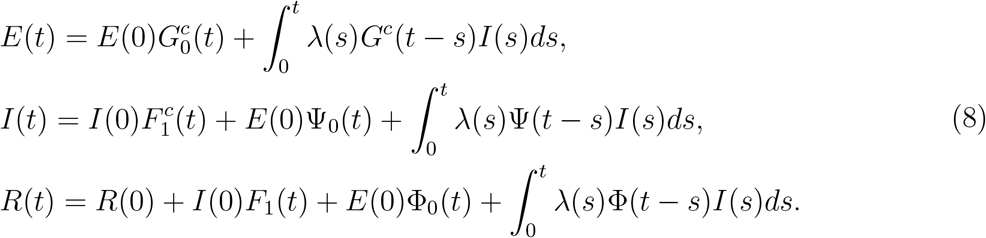

We shall assume that *E*(0), *I*(0) and *R*(0) are as in (5) and that 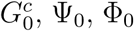 and *F*_1_ are given by (6), for some *ρ* > 0, and that the function *λ*(·) satisfies

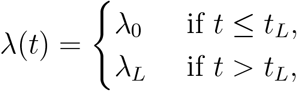

where *λ*_0_ satisfies (4) and *λ*_*L*_ satisfies the same equation with *ρ* replaced by *ρ*_*L*_ ≠ *ρ* (which may be negative).

Then, by Proposition 3, for all *t* ≤ *t*_*L*_,

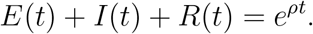

Moreover, by the second part of Proposition 3, we expect that there exists *δ* > 0 such that, for all *t* ≥ 0,

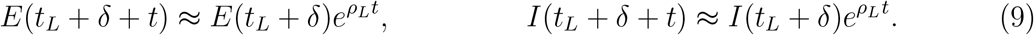

Summing the three equations in (8), we see that,

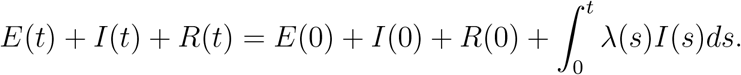

Let us write *EIR*(*t*) = *E*(*t*) + *I*(*t*) + *R*(*t*). As a result, if (9) holds, then, for all t ≥ 0,

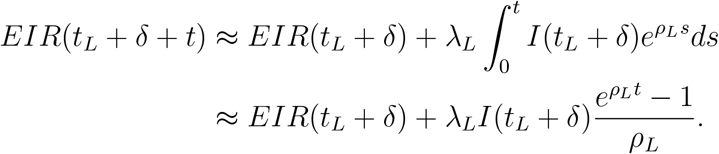

Moreover, by linearity, there exist positive constants *C*_*E*_(*δ*), *C*_*I*_(*δ*), *C*_*R*_(*δ*), depending only on *δ, ρ, ρ*_*L*_ and the distribution of (ℐ, *ε*), such that

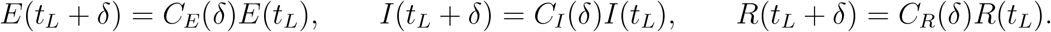

(We shall omit the dependence on *δ* in the following.) These constants are not known explicitly, but can be computed numerically by solving (8). Using this and Proposition 3, we obtain the following

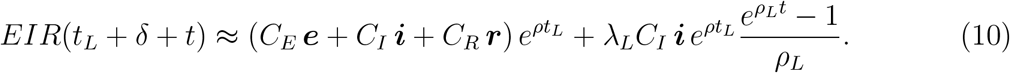

We shall use this later on to estimate the distributions of the delay between infection and hospital admission, hospital death and ICU admission.

### 1.4 Distribution of the sojourn times for Covid-19

The evolution of infectiousness for Covid-19 from the time of infection remains uncertain, but some early studies already provide constraints on the distribution of the sojourn times (*ε*, ℐ) for this disease. In particular, [HLW^+^20] estimate that infectiousness starts as early as 2.3 days before the onset of symptoms and declines within 7-8 days of symptom onset. Assuming that symptoms start on average 5.2 days after infection (as reported in [HLW^+^20]), we conclude that *ε* is between 2 and 4 days, and that ℐ is at least 3 days (if the infected individual is isolated shortly after the onset of symptoms) and no more than 10 to 12 days.

For the exposed time *ε*, we shall take either a fixed value of 3 days or a linear combination of the form 2 + 2 *X*_1_, where *X*_1_ is a symmetric Beta random variable with support in [0, 1].

In this study, we assume that infected individuals are divided in two groups. Individuals in the first group, called the *reported* individuals, are isolated shortly after the onset of symptoms, either because they self-isolate at home or because they are admitted to a hospital, where we assume that they do not infect anyone else. For these individuals, we assume that ℐ = 3 + *X*_2_, where *X*_2_ is a symmetric Beta random variable with support in [0, 1].

By contrast, individuals in the second group remain infectious for much longer, either because they show very mild symptoms or no symptoms at all or because they fail to be isolated. For these *unreported* individuals, we assume that ℐ = 8 + 4 *X*_3_, where *X*_3_ is a symmetric Beta random variable with support in [0, 1].

Let *p*_*R*_ be the proportion of reported individuals and let *Y* be a Bernoulli random variable with parameter *p*_*R*_, independent of *X*_2_ and *X*_3_. Then we can write

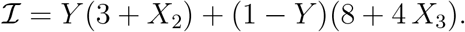

We thus see that the distribution of ℐ is bimodal, with a first peak at around 3.5 days and a second one around 10 days.

It seems that asymptomatic individuals are less infectious than symptomatic ones (see [CSC^+^20]). We could have included in the model different values of infectivity depending upon the duration of the infectious period of each individual, using the theory developed in [FPP20], but we lack quantitative information about the various levels of infectivity to make serious predictions, while the available data does not allow us to estimate many parameters. Note also that, as shown in [FPP20], during the early phase of the epidemic, the product of the number of asymptomatic patients and their infectivity determines the evolution of the epidemic. Overestimating the infectivity leads to an underestimation of the number of cases, which has no significant impact on the dynamic of the epidemic during the early phase, but may lead to underestimation of the proportion of the population which has acquired immunity at the end of this early phase.

### 1.5 Estimating the state of the epidemic prior to lockdown measures

In France, as of June 2020, the Covid-19 epidemic underwent three main stages. First, a rapid exponential growth (of the number of cases, number of hospitalized patients, number of deaths), followed by a slowing down of the epidemic due to the lockdown measures put into place around 16 March, settling after a few weeks to an exponential decrease of the number of newly hospitalized patients and new deaths. The third phase started on 11 May, with the progressive lifting of lockdown restrictions.

Since testing has been limited to a subset of symptomatic individuals and tests were performed at various intervals following symptom onset, the number of reported positive cases might not be exactly proportional to the true number of infected individuals throughout the different stages of the epidemic (and more importantly the ratio of infected individuals to tested individuals may vary significantly between the different phases). Hospital deaths, however, have been reported daily from 15 February. Moreover, from 18 March onwards, Santé Publique France has published daily reports of newly hospitalised patients, newly admissions in intensive care units (ICU) and new deaths in each administrative *département*, [San20].

Assuming that the distributions of the intervals between infection and hospital admission, admission in ICU and death do not vary over time, the growth rate of these quantities is necessarily identical to that of the cumulative number of infected individuals. As a result, if the distribution of the exposed and infectious periods (*ε*, ℐ) is known, we can infer the contact rate *λ* corresponding to a given growth rate *ρ* using (4).

Note that, since Santé Publique France only started publishing regional data on 18 March, the growth rate is inferred from the slope of the cumulative deaths during the first week of lockdown. Since the delay between infection and death in hospital is believed to be at least 10 days (*e.g*., [SKL^+^20]), it is safe to assume that all the individuals who died during this period were infected before lockdown measures took effect (however, see Subsection 2.1 below for the particular case of the Grand Est and Hauts-de-France regions).

If moreover one knows the distribution of the interval 𝒟 between infection and death, for example, as well as the infection fatality ratio *f* (the probability that an individual eventually dies, given that he or she has been infected), it is possible to infer the state of the epidemic. Indeed, the cumulative number of deaths at time *t* ≥ 0, denoted by Λ_*D*_(*t*), is then given by

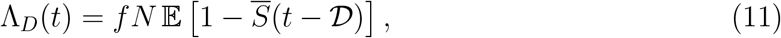

where *N* is the total population size (the expectation is taken with respect to the random variable 𝒟). During the early phase of the epidemic,

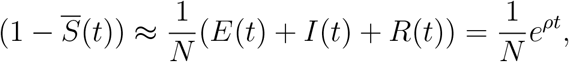

where *t* is the time elapsed since the (unknown) time of the start of the epidemic (*i.e*., the theoretical time at which only one individual was infected in the population), and *E*(*t*), *I*(*t*) and *R*(*t*) solve the linearized system (1). Thus, during this phase,

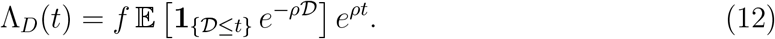

Hence, if one knows the infection fatality ratio and the distribution of 𝒟, one can infer both *ρ* and *t* from the observed cumulative number of deaths prior to lockdown measures. Using (4), one can deduce the value of the contact rate before lockdown measures, *λ*_0_, (given that the distribution of (*ε*, ℐ) is known) and thus simulate the epidemic up to this time.

Note that if *p*_*H*_ is the probability that a given infected individual is admitted to a hospital at some point, and if 𝒟_*H*_ is the interval between the time of infection and hospital admission, then, the cumulative number of hospitalized patients at time *t* ≥ 0, denoted by Λ_*H*_(*t*), is given by

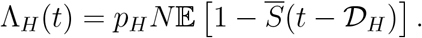

Hence, during the early phase of the epidemic,

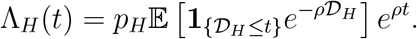

As a result, one can obtain the probability of being admitted to hospital if one knows *f* and the distributions of 𝒟 and 𝒟_*H*_ through the relation,

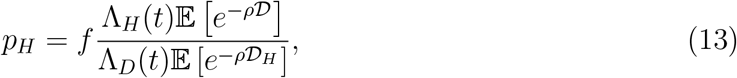

for any time *t* ∈ [0, *t*_*L*_] for which 𝒟 < *t* and 𝒟_*H*_ < *t* almost surely, where *t*_*L*_ is the time at which lockdown measures are put into place.

### 1.6 Estimating the contact rate during lockdown measures

At the start of lockdown measures, the contact rate in the population dropped sharply over the course of a few days. As a result, the daily number of deaths in hospitals started to slow down, before decreasing at a steady rate after approximately three weeks. In addition, the number of new hospital admissions and ICU admissions also decreased at a steady rate after a (shorter) transition period.

Assuming that, at this point, the proportion of susceptible individuals in the overall population remains close to 1, this corresponds to the situation described by (8) with *ρ*_*L*_ < 0. Using (10) and (11) and choosing *t* large enough (such that ℙ (𝒟 > *t*) is small), we have

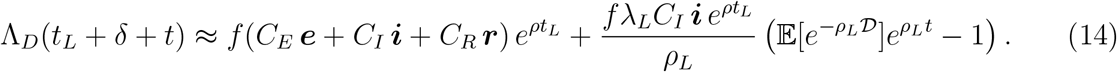

In particular, there exists a constant *C* > 0 such that

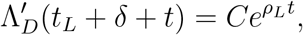

and the same holds for hospital admissions (with a different constant). As a result, we can estimate *ρ*_*L*_ from the cumulative number of hospital admissions and hospital deaths, and deduce the value of the contact rate during lockdown using (4).

Here and in what follows, we assume that the distribution of the sojourn times (*ε*, ℐ) is unaffected by the various containment measures put into place during the course of the epidemic. This may not be true, especially if contact tracing and massive testing are implemented at a sufficient scale in the population, but the extent of such measures in France, at least during the early stages of the Covid-19 epidemic, remained very limited.

### 1.7 Estimating the distribution of the delay between infection and hospital death or hospital admission

We have seen above that (14) can be used to determine the contact rate during lockdown measure, but in fact we are able to estimate all the constants appearing in (14) by fitting an exponential curve to the cumulative number of hospital deaths. More precisely, we can estimate the values of

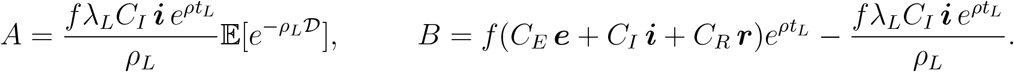

The only unknowns here are 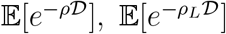 and *t*_*L*_ (since *t*_*L*_ is determined using the distribution of 𝒟 through (12)). Substituting (12) (with *t* = *t*_*L*_) in these equations, we can remove *t*_*L*_ and we obtain

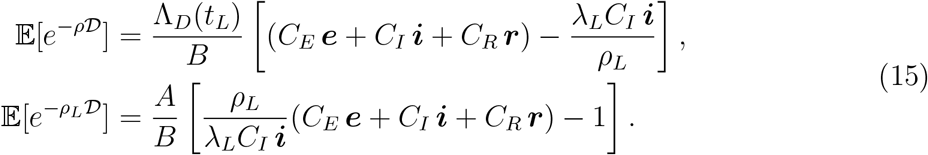

These two equations do not fully characterize the distribution of 𝒟, but if we assume that this distribution belongs to some two-parameter family, it is possible to estimate the corresponding parameters. In the following, we shall assume that 𝒟 follows a Gamma distribution with parameters (*θ, k*). Then, for all *ρ* > −*θ*^−1^,

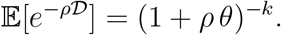

The parameters *θ* and *k* can then be computed numerically by inverting (15).

We proceed in the same way for the delay between infection and hospital admission and ICU admission. Once we have estimated the parameters of the delay distribution for each of these events, we can compute *t*_*L*_ using (12) and the probabilities of hospital admission and ICU admission using (13).

### 1.8 Estimating the contact rate after the easing of lockdown measures

The easing of lockdown restrictions was organized in different phases. On 11 May, people were allowed to leave their homes, shops started to reopen and schools progressively welcomed pupils again. On 2 June, bars and restaurants reopened in most of the country, and on 22 June, all schools reopened, along with cinemas, and most activities resumed, although sanitary measures continued to be enforced (*e.g*. wearing a mask remains mandatory in mainy public places including trains and public transports).

After the easing of lockdown restrictions, on May 11, we can assume that the contact rate in the population gradually shifted to a new value, *λ*_*E*_, corresponding to another growth rate *ρ*_*E*_. As we shall see below, at the end of the lockdown period, the proportion of susceptible individuals in the population had already dropped by 5 to 10%, at least in Î le de France and the Grand Est and Hauts-de-France regions. As a result, the approximation 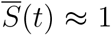 may not be valid at this point, preventing us from directly applying (4). Since the epidemic keeps decreasing, however, the value of 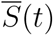 does not vary much during the period over which we estimate the new growth rate *ρ*_*E*_. It follows that we can replace *λ* by 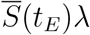 in (4), where 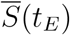 is the proportion of susceptible individuals at the end of the lockdown period, and hence deduce the corrected value of the contact rate.

The same correction should also be applied in the future when estimating the contact rate in later stages of the epidemic.

### 1.9 Estimating the state of the epidemic

Once we have estimated the contact rates prior to lockdown (*λ*_0_), during lockdown (*λ*_*L*_) and after lockdown (*λ*_*E*_), as well as the interval between the (theoretical) index case and the start of lockdown *t*_*L*_, we are in a position to compute the state of the epidemic. To do this we numerically solve the equations of Definition 1 with

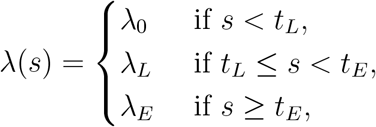

where *t*_*L*_ is time at which lockdown measures are implemented and *t*_*E*_ is the time at which these measures are eased. For the initial condition, using Proposition 3, we choose

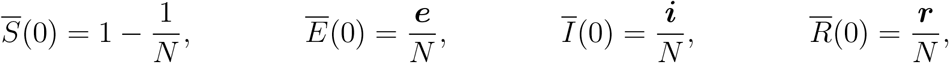

with ***e, i, r*** as in (1) and 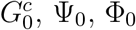 as in (6), choosing *ρ* equal to the growth rate prior to lockdown in (1) and as the parameter of the exponential random variable Θ in (6). In this way, by Proposition 3, 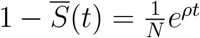 for all *t* ≤ *t*_*L*_, as expected.

It might seem counter-intuitive to start the epidemic with some fraction of removed individuals 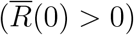, even more so as we assume that only one individual is not susceptible at this time. This, however, is only an artefact of our model. We are not trying to estimate the *true* initial state of the epidemic; we merely find a suitable initial condition so that the observed exponential growth prior to lockdown measures fits the observed data.

## 2 Results

For our estimation of the state of the Covid-19 epidemic in France, we split the country in three regions, or patches: the Grand Est and Hauts-de-France regions (the northeastern part of the country where the epidemic took off earliest), Î le de France (*i.e*. the densely populated Paris region) and the rest of the country, excluding Corsica and the overseas territories.

The exclusion of Corsica was motivated by its singular behaviour. Indeed, in this island, the epidemic took off very quickly in March, followed by a very sharp plateau, much more marked than in the rest of the country. In any case, including Corsica in our analysis should not significantly affect our results.

### 2.1 Growth rates of the epidemic

We measured the growth rate of the cumulative number of infected individuals during the early phase of the epidemic by fitting an exponential curve to the cumulative number of deaths in each patch, between 19 March and 26 March (earlier data was only available at the national level). Despite the fact that very strict lockdown measures were already effective at the time, hospital deaths continued to increase exponentially until at least 26 March, mainly due to the fact that infected individuals who died of Covid-19 during this period had been infected before national lockdown, hence during the exponential growth phase. The growth rates varied between 0.20 in the Grand Est and Hauts-de-France regions and 0.26, 0.28 in the rest of the country and in Î le de France, corresponding to a doubling time of the cumulative number of infected individuals of around 2.5 days (see Table 1).

**Table 1:** Growth rates measured prior to lockdown measures, in the steady phase of the lockdown period and after the easing of lockdown restrictions in each of the three patches used for the estimation (Î le de France, Grand Est and Hauts-de-France, and the rest of mainland France, excluding Corsica). Growth rates prior to lockdown are measured using hospital deaths between 19 March and 26 March (except for the whole country for which we used data from 1st March).

| Region | Île de France | Grand Est<br>and Hauts-de-France | Rest of<br>France (ex-<br>cluding<br>Corsica and<br>overseas) | Mainland<br>France |
| --- | --- | --- | --- | --- |
| Growth rate during the<br>early phase (from hospital<br>deaths) | 0.28 | 0.20 | 0.26 | 0.23 |
| Growth rate under lock-<br>down | -0.060 | -0.053 | -0.051 | -0.055 |
| Growth rate after 11 May | -0.046 | -0.057 | -0.052 | -0.052 |
| Growth rate after 2 June | -0.0028 | -0.033 | 0.016 | -0.0035 |

The relatively slow growth rate in the Grand-Est and Hauts-de-France regions might seem surprising, given that this is where the epidemic first took off at a very quick pace. We interpret this as the result of emergency measures which were put into place from 7 March in the most affected areas: school closures, banning of public events and gatherings, *etc*.[AFP20]. Indeed, if the epidemic had been growing at this rate of 0.20 from the beginning, our model shows that it should have started much earlier than it actually did, and the number of hospital deaths in the whole country would have grown more slowly (but for a longer time) at this time, as shown on Figure S1. If instead we assume that the growth rate in this region was 0.27 (*i.e*. the growth rate at the national level) up until 7 March, then 0.20 between 7 March and 16 March, the SEIR model predicts a cumulative number of hospital deaths much closer to what was actually observed at the national level (see Figure S1).

After a few weeks of lockdown, daily hospital admissions and deaths started to decline, again exponentially, at a steady rate in each patch, as shown on Figure 3. We observed a drop in the number of daily admissions and deaths every week-end, which was systematically compensated at the start of the following week, indicating reporting delays. We thus fitted simultaneously three exponential curves to the *cumulative* number of hospital admissions, hospital deaths and ICU admissions between 3 April and 11 May (starting only on 13 April for ICU admissions and deaths). The obtained values are displayed in the second line of Table 1. Overall, the growth rate of the epidemic in each patch stood between −0.051 and −0.06, corresponding to a halving time of the number of infected individuals of around 12 days.

**Figure 3:**
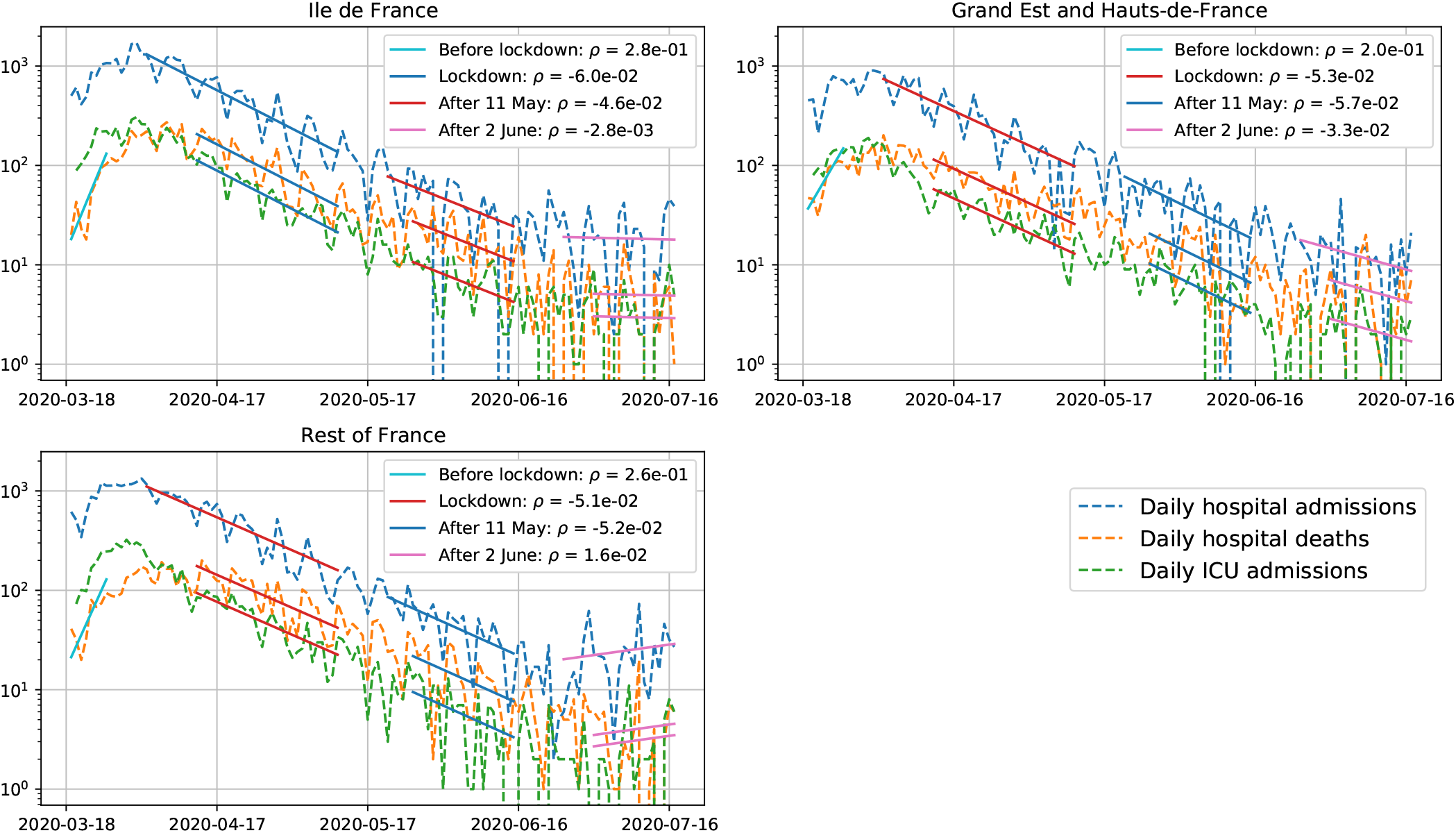
Growth rates of the cumulative number of infected individuals during (*ρ*_*L*_) and after (*ρ*_*E*_) lockdown in France, deduced from the number of hospital admissions, ICU admissions and hospital deaths in each patch. In each instance, an exponential curve was fitted simulateneously to each cumulative number of hospital admissions, ICU admissions and deaths. The last value in each patch is deduced from the cumulative number of hospital admissions, ICU admissions and hospital deaths during the last two weeks of June.

Using the same method, we can estimate the new growth rate of the cumulative number of infected individuals after the easing of lockdown measures, on 11 May. Overall, the epidemic appears to be decreasing but the decrease appears to slow down after the easing of lockdown restrictions in Î le de France (− 0.046 instead of −0.06 during lockdown), reflecting the increased contact rate in this region. This seems to be confirmed by the most recent data, which suggests an increased propagation of the virus in Î le de France and in the Grand Est and Hauts-de-France regions (third set of solid lines on Figure 3). This raises fears that further lifting of restrictions might cause this growth rate to increase even further, eventually triggering a second epidemic wave. Indeed, not enough time has elapsed since the last easing of restrictions (on 2 June and even later on 22 June) in order to accurately estimate the new contact rate in the population.

Note that the SEIR model predicts a small but non-negligible (of the order of 5 to 10%) level of acquired immunity, at least in Î le de France and the Grand Est and Hauts-de-France regions (see Subsection 2.3). We would thus expect to observe a progressive acceleration of the exponential decrease in the number of new hospital admissions and deaths. The fact that this does not take place suggests that the easing of lockdown restrictions somewhat compensates the effect of this acquired immunity.

Since the end of June, daily hospital admissions, ICU admissions and hospital deaths seem to stagnate at fairly low levels, and even to increase slightly in some regions. This could be due to the fact that the effective reproduction number in France is very close to one, or could be the sign of the start of a second epidemic wave. Note however that, because of the delay between infection and hospital admission (which we estimate below to be around 14 days on average), our estimate of the contact rate is always at least two to three weeks behind the current one, and the true number of infected individuals could have been increasing for the past two weeks without affecting the observed hospital admissions.

It is worth mentioning the fact that other datasets seem to indicate an increase in the number of cases over the first weeks of July 2020. This is the case of the Covid-19-related actions reported by SOS Médecins, which performs emergency medical interventions around the clock in every part of the country. Including this dataset in the estimation of the growth rates does not significantly change the values obtained during and just after lockdown, but yields positive growth rates after mid-June (respectively 0.042 in Î le de France, 0.057 in the Grand Est and Hauts-de-France regions and 0.068 in the rest of the country, corresponding to a doubling time of between 4.5 and 7 days), as shown in Figure S2.

### 2.2 Reproduction numbers and the effect of lockdown measures

Using the relation (4) and our assumptions on the distribution of (*ε*, ℐ) from Subsection 1.4, we can estimate the values of the reproduction number *R*_0_ in each patch during each phase of the epidemic. Since the growth rates prior to lockdown and during lockdown are relatively uniform across the different patches (respectively, around 0.27 and −0.049, taking into account our interpretation of the lower growth rate in the Grand Est and Hauts-de-France regions), we use the same value for all patches.

The proportion of reported individuals affects both the expectation of the infectious period 𝔼 [ℐ] and the whole distribution of (*ε*, ℐ). Considering Corollary 4, we thus observe that the estimated value of the reproduction number depends on the proportion of reported individuals, and more generally on the whole distribution of (*ε*, ℐ).

If we assume that all infected individuals are reported (*p*_*R*_ = 1), the reproduction number in France is 3.4 prior to lockdown measures and 0.79 during lockdown. If, however, we assume that all individuals are unreported (*p*_*R*_ = 0), the reproduction number in France is 6.3 prior to lockdown and 0.66 during lockdown. The estimate of the reproduction number then varies continuously between these values for intermediate values of the proportion of reported individuals. For instance, if the proportion of reported individuals is close to 0.8, the reproduction number is estimated to be 4.2 during the early phase of the epidemic and 0.73 during lockdown, whereas if this proportion is 0.2, *R*_0_ = 6 before lockdown and 0.67 during lockdown. Note that our estimate of *R*_0_ fits well the results in [SLX^+^20].

The relatively large values (around 6) obtained for small proportions of reported individuals suggest that this proportion is closer to 0.8 than to 0.2. Moreover, the values we obtain, even for large proportions of reported individuals, are sometimes larger than other estimates in the literature (Table 2). We can attribute this to two things. First, as shown in Figure 2, the Markovian SEIR model tends to slightly underestimate *R*_0_ in the two regimes we are interested in (growth in the early phase and decrease under lockdown). Second, some studies do not take into account the exposed period *ε* in their model. However, Corollary 4 shows that neglecting the exposed period leads to underestimating of the reproduction number *R*_0_, here by a factor of 0.76 for each day of exposed period (*e*^−*ρ*^ = 0.76 with *ρ* = 0.27). Finally, uncertainty about the infectious period also ℐaffects the estimates of the reproduction number.

**Table 2:**
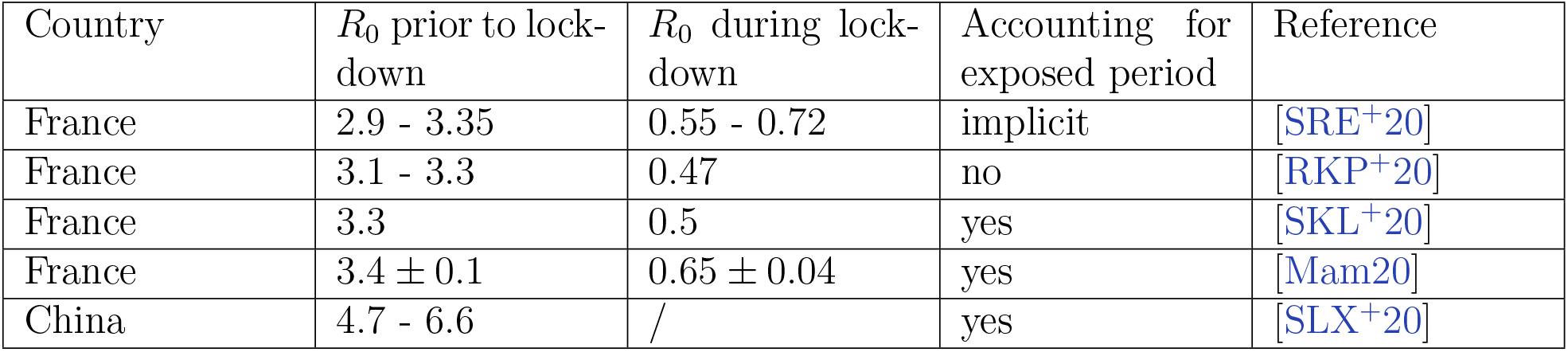
Estimates of the reproduction number in the literature

| Country | $R_0$ prior to lockdown | $R_0$ during lockdown | Accounting for exposed period | Reference |
| --- | --- | --- | --- | --- |
| France | 2.9 - 3.35 | 0.55 - 0.72 | implicit | [SRE<sup>+</sup>20] |
| France | 3.1 - 3.3 | 0.47 | no | [RKP<sup>+</sup>20] |
| France | 3.3 | 0.5 | yes | [SKL<sup>+</sup>20] |
| France | $3.4 \pm 0.1$ | $0.65 \pm 0.04$ | yes | [Mam20] |
| China | 4.7 - 6.6 | / | yes | [SLX<sup>+</sup>20] |

### 2.3 State of the epidemic and acquired immunity at the population level

Using (12) and Subsection 1.9, we can estimate the state of the epidemic in each patch, given the infection fatality ratio *f* and the distribution of the delay 𝒟. Unfortunately, these two quantities are notoriously hard to measure during the early stage of the epidemic.

The infection fatality ratio for the Covid-19 epidemic in France has been estimated in at least two studies [RKP^+^20, SKL^+^20]. Both studies found a fatality ratio of 0.5%, with significant variation across age classes in [SKL^+^20]. These studies only account for hospital deaths, as do we, even though a significant number of deaths take place outside hospitals, mainly in nursing homes. Hence this ratio has to be corrected (roughly by a factor 1.6 [RKP^+^20]) to obtain the true infection fatality ratio of Covid-19 (and to obtain correct predictions for the expected number of deaths). Nevertheless, since we use hospital deaths to calibrate our SEIR model, we shall use the infection fatality ratio estimated by [SKL^+^20, RKP^+^20] when using (12).

We can at least bound this ratio from below using the observed excess mortality in some regions. For example, in Lombardy (Italy), 13,575 people died of Covid-19, for a total population of 10 million, showing that this ratio is at least 0.14% (or 0.088% for hospital deaths). Another estimate of this ratio was obtained in a small town in Germany, where around 1,000 individuals were tested, out of which 15.5% tested positive and 7 people died, yielding an infection fatality ratio of 0.37% [SSK^+^20] (corresponding to 0.23% for hospital deaths).

On the other hand, the infection fatality ratio can be bounded from above by the apparent death rate, that is to say, the ratio of Covid-19 related deaths and declared positive cases, at least while the epidemic seems to be receding, as is the case in France in June 2020, with less than 50 deaths per day. Taking only hospital deaths into account, this suggests that *f* is no more than 12%. Other countries have much lower apparent death rates, *e.g*. South Korea (2.3%), Germany (4.6%). These discrepancies can mostly be attributed to differences in testing capacities and to a lesser extent to differences in hospital capacities.

Using the method described in Subsection 1.7, we estimated the parameters (*k, θ*) for the delay between infection and each of the three possible events (hospital admission, ICU admission and hospital death). The means and standard deviations of the distributions we obtained are given in Table 3.

**Table 3:**
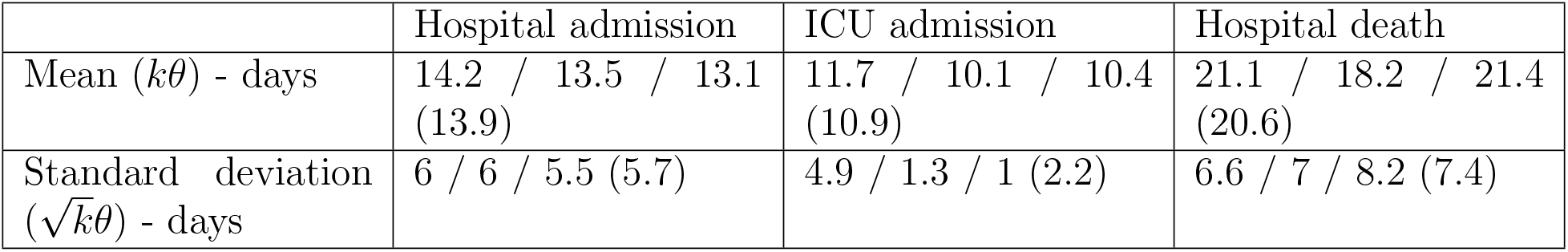
Mean and standard deviations of the delays between infection and hospital admission, ICU admission and hospital deaths obtained from (15). Each cell contains the values estimated for each of the three patches (Î le de France, Grand Est and Hauts-de-France and the rest of the country), plus in brackets the value obtained when considering a single patch for the whole country. The model assumed *p*_*R*_ = 0.8, but this parameter did not have a substantial impact on the results (*e.g*., assuming that *p*_*R*_ = 0 leads to 14.7 days on average between infection and hospital admission, 11.7 days before ICU admission and 21.4 days before hospital death).

We estimated the probabilities of being admitted to hospital given infection. These obviously depend on the infection fatality ratio, but, on the other hand, the death to admission ratio (*i.e*., the probability of dying given that one is admitted to a hospital) is constant. We estimated that the death to admission ratio was 0.19 in Î le de France, 0.20 in the Grand Est and Hauts-de-France regions and 0.16 elsewhere. Note that we consider that these do not evolve over time, which may not be the case, especially if hospitals become overwhelmed by the influx of patients. This could for example explain the discrepancy between the death to admission ratios in the different regions.

Figure 4 shows the predicted levels of immunity (*i.e*., the proportion of infected individuals) up to 31 July in each patch for three values of the infection fatality ratio *f*, using the delay distributions estimated above. Unsurprisingly, higher values of this ratio lead to lower predicted levels of immunity. Note that underestimating 𝒟 has the same effect as overestimating the infection fatality ratio, as can be noted from (12). As a result it is equally important to estimate *f* and the distribution of 𝒟, a fact which is too easily overlooked.

**Figure 4:**
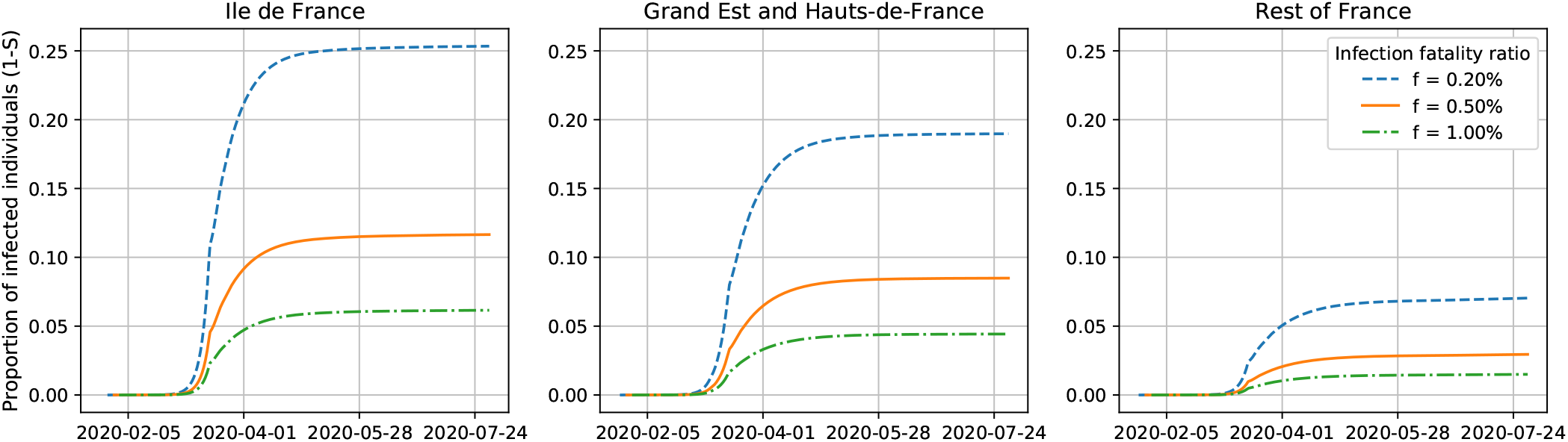
Predicted level of immunity in each patch as a function of the infection fatality ratio *f*. All the simulations use *p*_*R*_ = 0.8. The rightmost point of each curve corresponds to the value on 31 July

For the fatality ratio estimated by [RKP^+^20, SKL^+^20] (*f* = 0.5%), we estimate that between 8 and 10% of the population has been infected by mid June in the most affected regions (Î le de France, Grand Est and Hauts-de-France), and around 2 to 3% of the population in the rest of mainland France. As shown by Figure 4, these estimates vary significantly as a function of *f* and the distribution of the delay between infection and death (𝒟), highlighting the need for more accurate measures of these quantities.

## 3 Discussion

Our goal in this article is to study, in the context of an ongoing epidemic, what knowledge about the parameters of a given model can be extracted from a certain type of data, and what needs to be estimated separately. In addition, our aim is to stress how the choice of the model (for instance choosing a Markovian or a non-Markovian model) affects the predictions one can make based on the data.

As exposed in Remark 5, knowing the distribution of the sojourn times in each compartment (*ε*, ℐ) is crucial to estimate the value of *R*_0_ from the initial growth rate of the epidemic. Without knowledge on these sojourn times, the reproduction number can only be estimated using contact tracing, which is both difficult and costly. As shown in Figure 2, Markovian epidemic models (but not only) can in some regime underestimate the value of *R*_0_ for a given initial growth rate of the epidemic. If this is the case, then they will underestimate the final proportion of infected individuals in the population in the absence of containment measures, and thus the predicted death toll of the epidemic.

On the other hand, one needs additional information on the infection fatality ratio (*i.e*., the proportion of infected individuals who die eventually) and on the distribution of the delay between infection and death in order to estimate the true state of the epidemic at any given time and thus the level of acquired immunity in the population. We have shown that some information of this delay distribution can be retrieved from the time series of hospital deaths caused by the disease, but this information is partly contingent on the model used to estimate the state of the epidemic.

More crucially, one has to wait for some sort of containment measures to take effect in order to estimate these delays, hence this cannot be done during the initial growth phase of the epidemic. Since Markovian models tend to underestimate the inertia of the epidemic after containment measures are put into place (*i.e*., they tend to decrease exponentially too soon after a change in the contact rate), using these will lead to an underestimation of the length and height of the epidemic *plateau* reached before the decreasing phase. This can be seen in the slow decrease in hospital admissions and deaths even after 3 weeks of strict lockdown, something which the Markovian model would not have predicted. One way to force the Markovian model to display this kind of behavior would be to make the change in the contact rate more gradual, but we do not believe that this would reflect what took place in France. Another way to force the Markovian model to fit this behavior would be to overestimate the delay between infection and hospital death, which would artificially increase the Markovian model’s inertia, but this would also lead to an overestimation of *t*_*L*_, and hence of the cumulative number of infected individuals.

One thing that we can note is that the kind of data studied in this article carries very little information on which model is closer to reality, at least during the early stages of an epidemic. External information on the true delay distribution between infection and death, or infection and hospital admission, *etc*., can help to put constraints on the model, yielding more accurate predictions of the state of the epidemic.

The most recent data presents a few signs that the epidemic might be on the rise in different parts of the country, although the trend has yet to be confirmed. One indication of this is the fact that our model seems to underestimate the observed number of daily hospital admissions over the first two weeks of July, as can be seen on Figure 5. The SOS Médecins dataset is another sign that the growth rate of the epidemic might have been positive for a few weeks. The fact that hospital admissions, ICU admissions and hospital deaths are not increasing at such a steady rate yet could be due to the longer delay between infection and these events. Indeed, applying the method described in Subsection 1.7, we estimate that the average delay between infection and a call to SOS Médecins is around 8 days, much shorter that the 14 days estimated for hospital admission. If this trend is confirmed over the coming weeks by an increase in hospital admissions, then we can expect a second epidemic wave in France before the fall, as shown on Figure S3.

**Figure 5:**
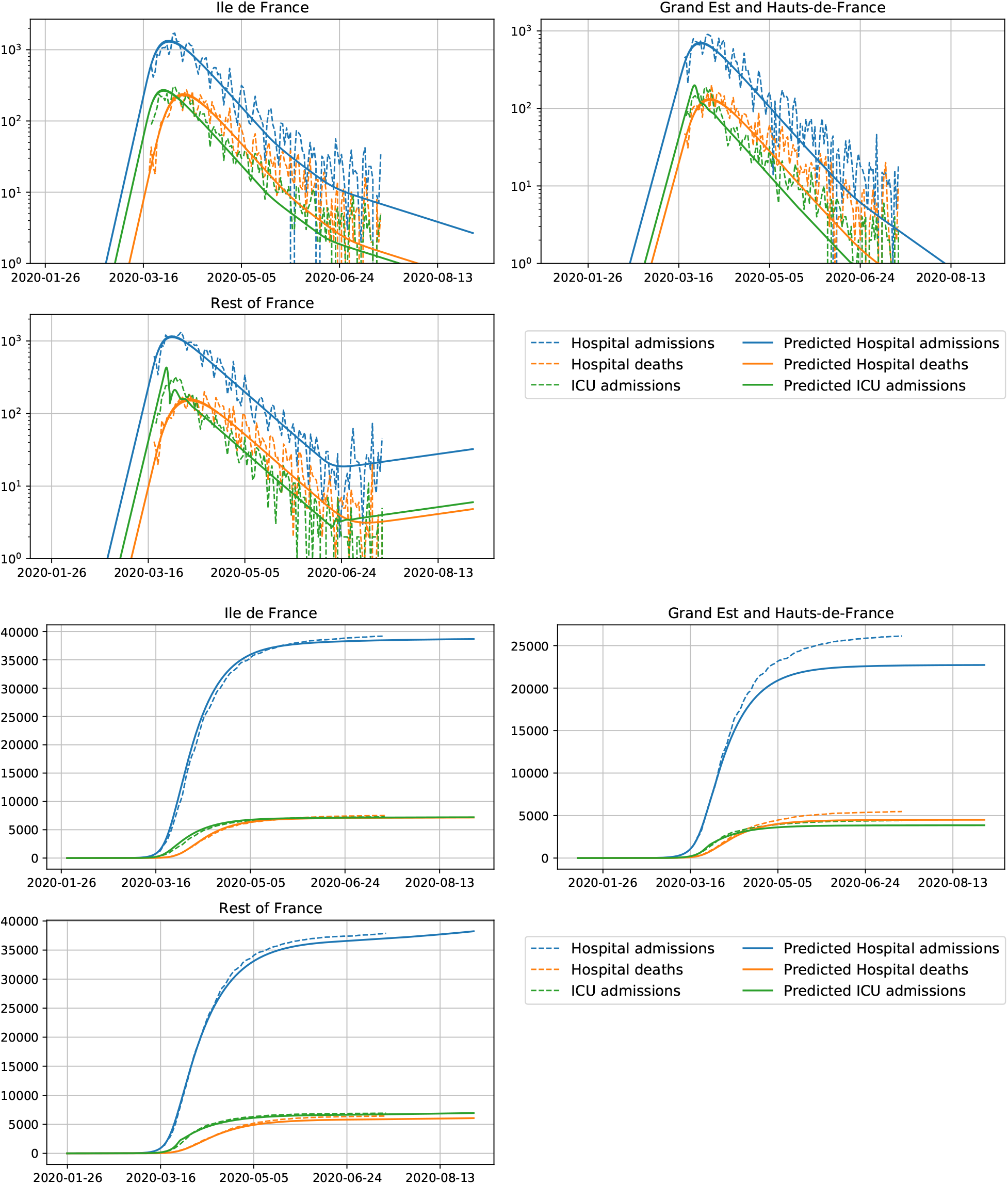
Observed hospital admissions, ICU admissions and hospital deaths in each patch (dashed lines) and the corresponding predictions (solid lines) of our calibrated non-Markovian SEIR model, up to 31 August 2020. Top graphics show the daily values, in logscale, while bottom graphics show the cumulative values in linear scale.

Note that an analysis similar to the one conducted in this paper can be carried out for a model where the infectivity of each individual is a random function of the time since infection, those random functions for the various individuals being i.i.d., see [FPP20]. This would yield a model which could account for the variability, both in time and between individuals, of the infectivity of each infected individual, something which is apparent for Covid-19 for example in [HLW^+^20]. Applying this type of models to the ongoing Covid-19 epidemic will be the subject of future work.

## Data Availability

The data used in this article is made public by Santé Publique France and is freely available online.

https://www.data.gouv.fr/fr/datasets/donnees-hospitalieres-relatives-a-lepidemie-de-covid-19/

## Code availability

The code used to simulate the SEIR model and to analyse the data is made available at https://github.com/rforien/Fit_Covid19_nonMarkovian.git

## A Proof of Proposition 3

We start with the case *ρ* > 0. Replacing (*E*(*t*), *I*(*t*), *R*(*t*)) in (2) by their expressions in (3) yields the following equations

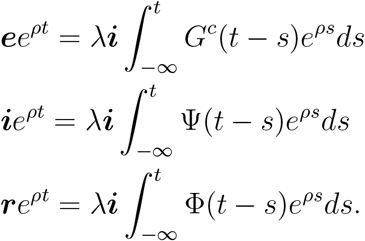

The second equation translates into

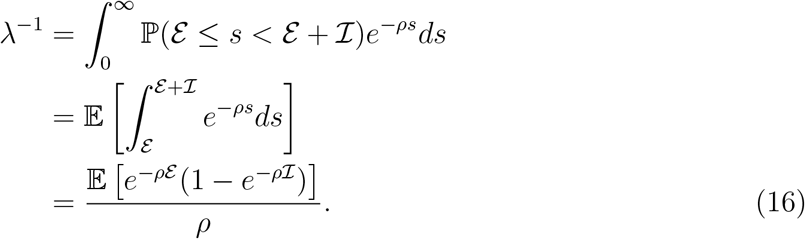

yielding (4). The remaining two equations give the relations between ***e, r*** and ***i***. The first one yields

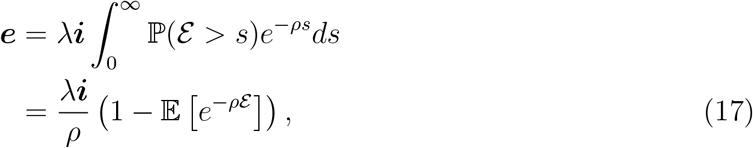

and the third one gives, for *ρ* > 0,

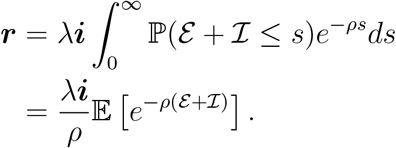

Combining these with the constraint ***e*** + ***i*** + ***r*** = 1 yields (5). Note that *λ****i*** = *ρ*.

To prove the second part of the statement, we only need to check that the quantities defined in (6) satisfy

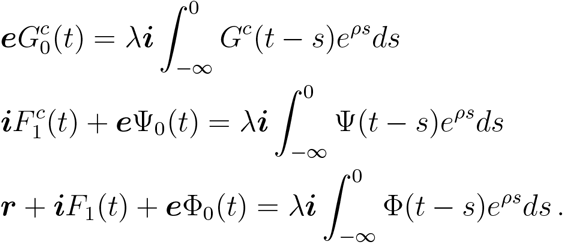

Indeed, combining these with (2) and the first part of the proposition, we obtain the fact that (3) solves (1). To check the first identity, write

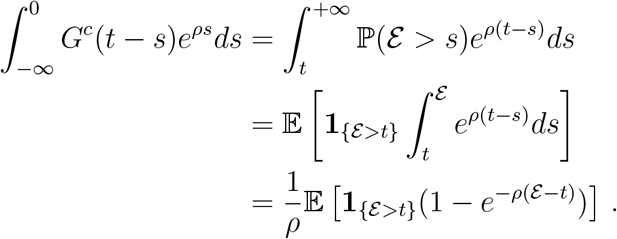

Multiplying by *λ****i***/***e***, we obtain

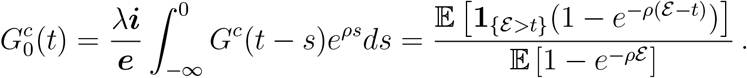

We conclude by noting that the term on right-hand-side equals ℙ (*ε* −Θ > *t* |Θ < *ε*), where Θ is an independent exponential random variable with parameter *ρ*.

Plugging (6) and (7) in the second equation, we obtain

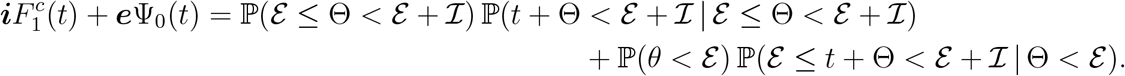

Since, for *t* ≥ 0,

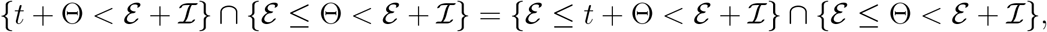

and

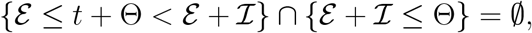

by the law of total probability,

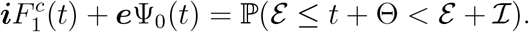

On the other hand, since *λ****i*** = *ρ*,

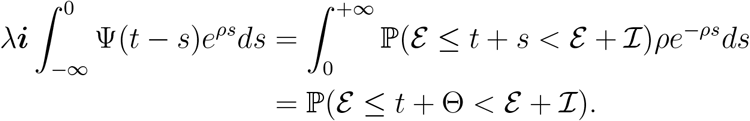

For the third equation, we proceed in the same way, plugging (6) and (7), we obtain

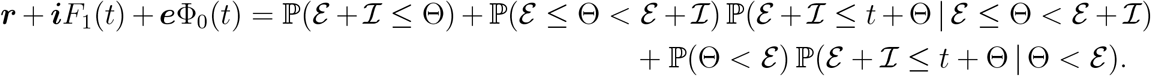

Now, we note that, for *t* ≥ 0,

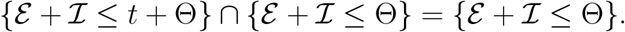

As a result, applying the law of total probability, we obtain

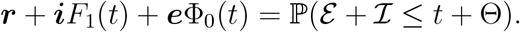

On the other hand, since *λ****i*** = *ρ*,

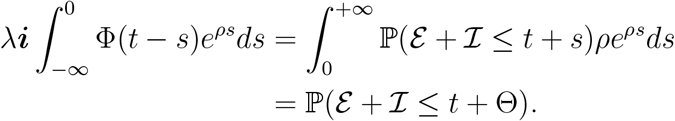

This concludes the proof of the first part of Proposition 3.

In the case *ρ* < 0, note that the computations in (16) and (17) remain valid, yielding the second part of the statement.

## B Including two-step measures in Grand Est and Hautsde-France

**Figure S1:**
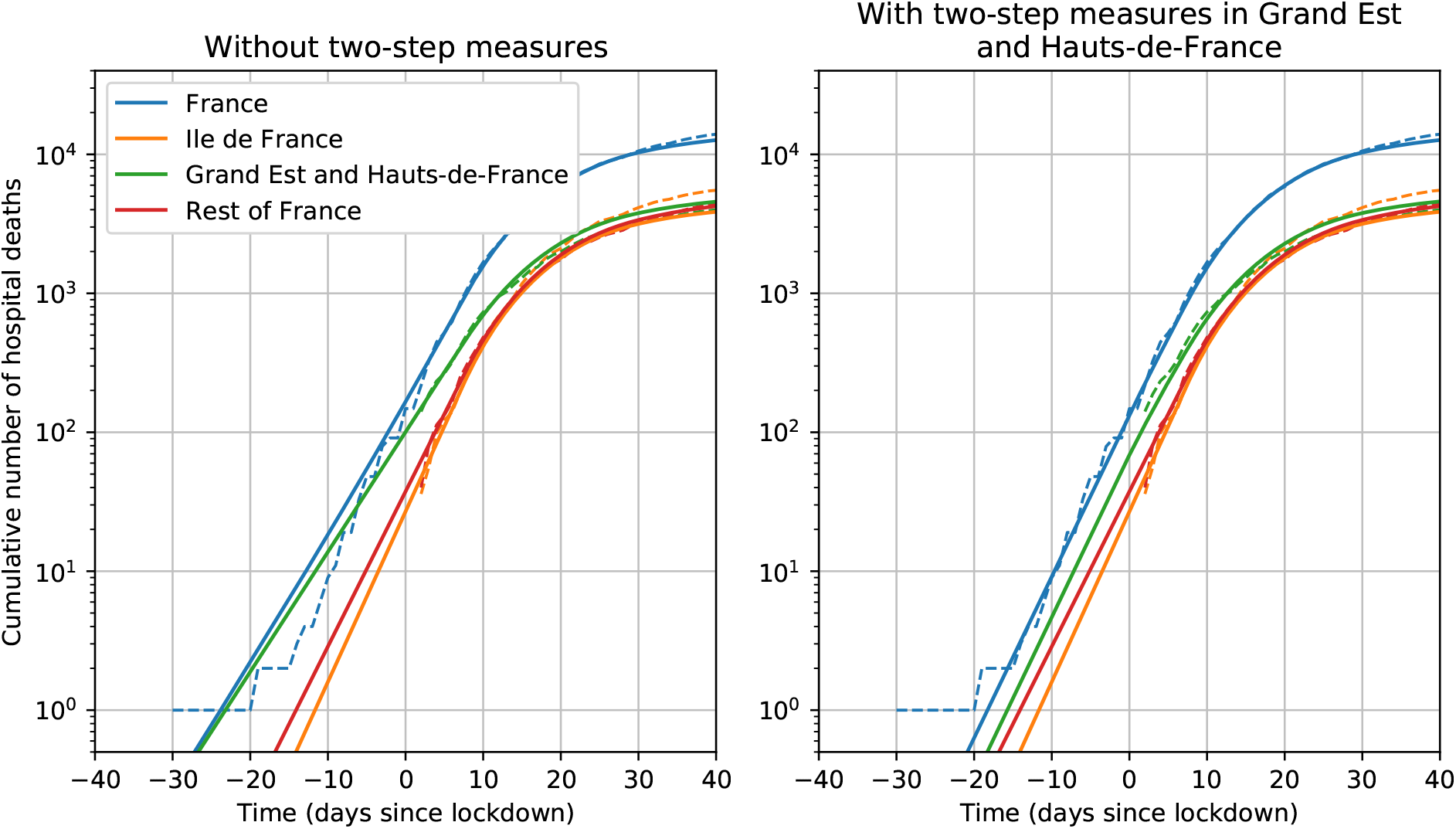
Predicted deaths during the early phase of the epidemic with and without two step measures in the Grand Est and Hauts-de-France regions. On the left, the growth rate inferred from the cumulative number of deaths in this patch is taken to be the growth rate of the epidemic in this region since the beginning, leading to a slow growth of the epidemic during the early phase, which does not fit the observations. On the right, the growth rate of the epidemic is 0.27 at first, and then 0.20 from 7 March, before changing again at the time of lockdown. The agreement with the observations is considerably improved.

## C Including the SOS M é decins dataset

**Figure S2:**
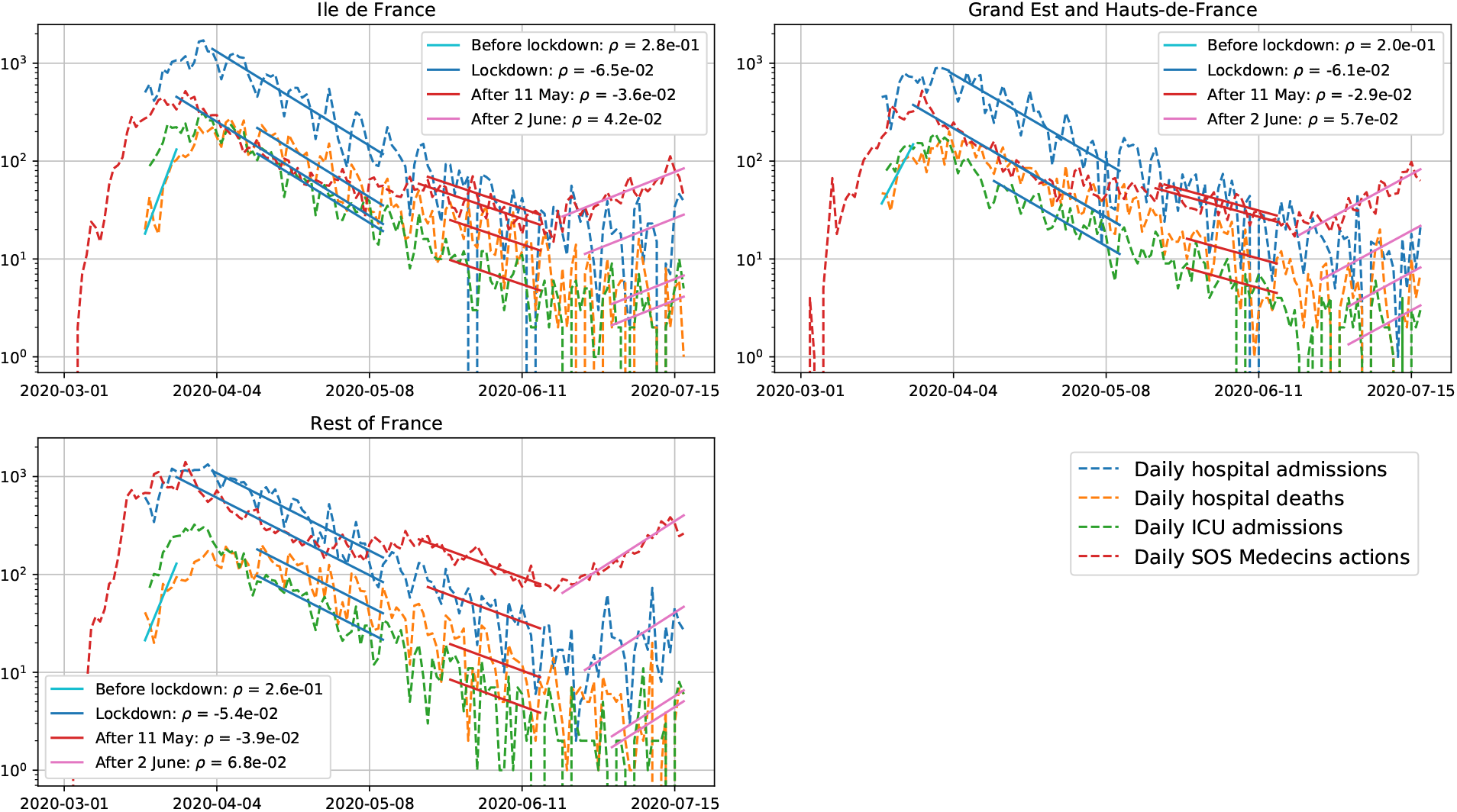
Estimation of the growth rates of the epidemic including the data from SOS M é decins. Including this dataset mainly affects the values obtained over the last observed period, suggesting that a second epidemic wave could be building up across the country.

**Figure S3:**
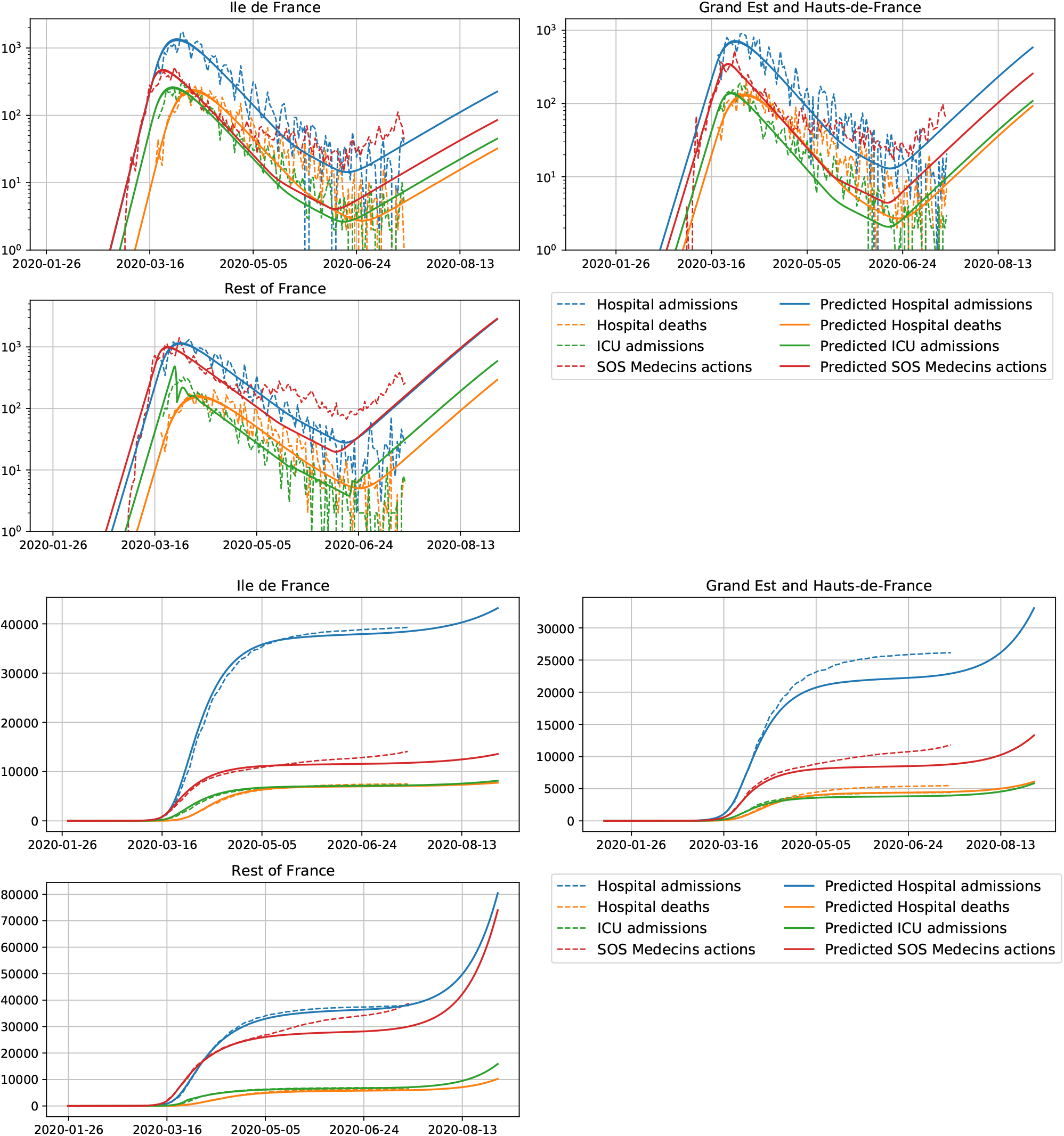
Prediction of the calibrated SEIR non-Markovian model accounting for the growth rates estimated using the SOS M é decins dataset. The predictions correspond to a second epidemic wave which reaches pre-lockdown levels sometime in September. Note that the prediction of the daily SOS M é decins actions is substantially off after 11 May 2020, which could be due to a change in the proportion of infected individuals who call this service around this time.

